# Estimating the cumulative incidence of SARS-CoV-2 infection and the infection fatality ratio in light of waning antibodies

**DOI:** 10.1101/2020.11.13.20231266

**Authors:** Kayoko Shioda, Max SY Lau, Alicia NM Kraay, Kristin N Nelson, Aaron J Siegler, Patrick S Sullivan, Matthew H Collins, Joshua S Weitz, Benjamin A Lopman

## Abstract

**Background:** Serology tests can identify previous infections and facilitate estimation of the number of total infections. However, immunoglobulins targeting severe acute respiratory syndrome coronavirus 2 (SARS-CoV-2) have been reported to wane below the detectable level of serological assays. We estimate the cumulative incidence of SARS-CoV-2 infection from serology studies, accounting for expected levels of antibody acquisition (seroconversion) and waning (seroreversion), and apply this framework using data from New York City (NYC) and Connecticut.

**Methods:** We estimated time from seroconversion to seroreversion and infection fatality ratio (IFR) using mortality data from March-October 2020 and population-level cross-sectional seroprevalence data from April-August 2020 in NYC and Connecticut. We then estimated the daily seroprevalence and cumulative incidence of SARS-CoV-2 infection.

**Findings:** The estimated average time from seroconversion to seroreversion was 3-4 months. The estimated IFR was 1.1% (95% credible interval: 1.0-1.2%) in NYC and 1.4% (1.1-1.7%) in Connecticut. The estimated daily seroprevalence declined after a peak in the spring. The estimated cumulative incidence reached 26.8% (24.2-29.7%) and 8.8% (7.1-11.3%) at the end of September in NYC and Connecticut, higher than maximum seroprevalence measures (22.1% and 6.1%), respectively.

**Interpretation:** The cumulative incidence of SARS-CoV-2 infection is underestimated using cross-sectional serology data without adjustment for waning antibodies. Our approach can help quantify the magnitude of underestimation and adjust estimates for waning antibodies.

**Funding:** This study was supported by the US National Science Foundation and the National Institute of Allergy and Infectious Diseases.

## Introduction

Severe acute respiratory syndrome coronavirus 2 (SARS-CoV-2), the virus that causes Coronavirus Disease 2019 (COVID-19), rapidly spread across the world in 2020.^1^ Globally, there have been 36 million documented cases and more than one million COVID-19 associated fatalities as of October 15, 2020, with case counts continuing to increase.^2^ Reliable measurement of infection history in a population is a critical epidemiologic outcome, and is needed to derive several key epidemiologic indices such as the infection fatality ratio (IFR). However, the number of SARS-CoV-2 infections reported through public health surveillance mechanisms is underestimated because of the limited capacity of testing and surveillance systems, an overwhelmed healthcare system, low healthcare-seeking behavior among those with mild disease, imperfect sensitivity of diagnostic tests (especially rapid antigen tests) and a large fraction of infections that are asymtomatic.^3^ Assays that detect viral antigen or genomic material cannot identify individuals who were previously infected once viral material is no longer present. In contrast, serology tests measuring the level of immunoglobulins have the potential to identify those previously infected. Serology results can be used to generate an estimate of the cumulative incidence, but this requires the relatively strong assumption that antibodies persist permanently after infection. Because antibody levels for SARS-CoV-2 wane over time and can become undetectable, some previously infected individuals could have already returned to a seronegative status at the time of testing (i.e. seroreversion). Hence, new methods are needed to account for waning antibodies when using cross-sectional serology data to estimate cumulative incidence of SARS-CoV-2 infection. The adjusted cumulative incidence can then be used to obtain more accurate estimates of critical epidemiologic measures, such as the IFR and the case ascertainment ratio.

Population-level serosurveys for SARS-CoV-2 have been conducted across the world, with variation in geographic scale, sample demographics, sampling mechanisms, and testing methods.^4,5^ Although the reported incidence of COVID-19 varies widely by location, a consistent finding in all settings is that total estimated infections vastly outnumber confirmed cases. For example, the Centers for Disease Control and Prevention (CDC) and commercial laboratories conducted large-scale geographic longitudinal serosurveys in 10 sites in the United States in the spring and summer of 2020.^6^ Seroprevalence ranged from 1% to 6.9% across sites. That study estimated that the number of total infections was 6-24 times higher than that of documented cases, while acknowledging that these ratios varied widely depending on the timing of sampling or the stage of the epidemic in each location. These early population-level serosurveys provided critical insights on the true burden of COVID-19; however, as we enter the 10th month of the US epidemic, the ability to estimate the cumulative incidence directly from serosurveys is increasingly limited because antibody levels continue to wane and the corresponding serologic ‘record’ of historical infection is lost. Indeed, Ibarrondo *et al*. estimated that the half-life of anti-SARS-CoV-2 spike receptor-binding domain IgG was 36 days.^7^ Patel *et al*. reported that over half (11/19) of health care personnel who tested seropositive in early April became seronegative at a second visit in June (approximately 60 days after the baseline).^8^ Taken together, these findings suggest that serosurveys have likely failed to recognize previous infection in those whose antibody levels have already waned below the detectable limit at the time of sampling. An adjustment for waning antibody kinetics must account for the interacting timescales of antibody kinetics, case incidence and the period over which a serosurvey is conducted.

Here, we describe a framework for estimating the cumulative incidence and IFR of SARS-CoV-2 from population-level cross-sectional serology data and mortality data, by adjusting for the timeline of seroconversion (acquisition of the detectable level of antibodies) and seroreversion (loss of detectable antibodies). We apply this framework to data from New York City and Connecticut, because these two sites observed a large wave of COVID-19 cases that lasted for a relatively short period in the spring 2020, followed by low case counts in the summer and early fall (Figure S1). We note that it is critical to distinguish the ability to detect antibodies from immunity to reinfection, which may persist longer than antibodies are detectable.^9^ We make no claim that the time from seroconversion to seroreversion estimated from this framework reflects the duration of immunity protection against reinfection.

## Methods

### Population-level cross-sectional seroprevalence data

Details of the serosurvey conducted by the CDC and commercial laboratories can be found elsewhere.^6,10^ Briefly, the survey collected blood samples in 10 US sites (Connecticut, Louisiana, Minneapolis-St Paul-St Cloud metro area, Missouri, New York City metro area, Philadelphia metro area, San Francisco Bay area, South Florida, Utah, and Western Washington State) from March to July 2020, and expanded the serosurvey to all 50 states in August. Blood specimens were originally collected for reasons unrelated to COVID-19, such as for routine medical care or sick visits. Multiple rounds of surveys have been conducted at each site, approximately every 3-4 weeks (Table S1). Each round tested approximately 1,800 samples from each site.^11^ Samples were tested by an enzyme-linked immunosorbent assay (ELISA) against the SARS-CoV-2 spike protein that detects the total immunoglobulin response (IgA, IgM, and IgG) with sensitivity 96% (95% confidence interval (CI): 98.3-99.9%) and specificity 99.3% (95% CI: 98.3-99.9%).^6^ Publicly available seroprevalence data adjusted for age and sex distributions were downloaded on October 8, 2020.^12^ We focused our analysis on New York City and Connecticut because the single, short wave of infection in the spring allowed us to evaluate how the antibodies acquired over a short period subsequently waned during a time when minimal new infections were occurring in these locations (Figure S1). More details can be found in the supplementary methods.

### Mortality data and case data

Mortality data are less subject to changes in testing capacity and guidelines over time than case data, so we used mortality data to estimate parameters. We analyzed daily time series data on COVID-19 associated deaths from March to September 2020 in New York City and Connecticut. New York City data and Connecticut data were downloaded from their government websites on October 2 and 6, 2020, respectively.^13,14^ We used the total (i.e., probable and confirmed) deaths for our mortality time series (supplementary methods). To account for a delay distribution between symptom onset and death, we used data on the date of symptom onset and date of death for 6,999 COVID-19 associated deaths in Georgia, USA from February to October 2020.

To estimate case ascertainment ratios (see the “Model” section for detail), we also utilized daily time series data for the number of documented cases in New York City and Connecticut from their government websites (supplementary methods).^13,14^

### Model

We estimated the IFR and time from seroconversion to seroreversion using the reported number of deaths and CDC serosurvey data (Figure 1). Using a Markov chain Monte Carlo (MCMC) analysis, we estimated the number of seropositive individuals on day *t* (*S*_*t*_) as follows:

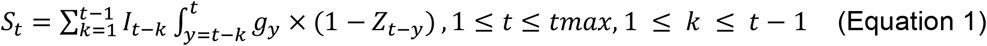

where *I*_*t*_ is the total number of SARS-CoV-2 infections on day *t*, which is the number of reported deaths divided by the estimated IFR, after accounting for the delay between symptom onset and death (supplementary methods). We estimated the IFR, assuming that the IFR was constant over time in the main analysis and relaxed this assumption in a sensitivity analysis (see the “sensitivity analysis” section for details). Additional parameters include *tmax*, the total number of days in the daily time series data for COVID-19 deaths, and *g*, the probability density function of the Weibull distribution for time from symptom onset to seroconversion with the mean 11.5 days and standard deviation 5.7 days,^15^ which is consistent with other reports.^16–19^ For asymptomatic cases, *g* represents the time from onset of infectiousness to seroconversion (supplementary methods). *Z* is the cumulative density function of the Weibull distribution for time from seroconversion to seroreversion. Therefore, 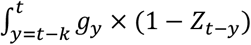 in Equation 1 represents the probability that an individual seroconverted before day *t* and seroreverted after day *t*, which in other words means the probability that an individual remains seropositive on day *t*. We estimated the mean of the Weibull distribution for time from seroconversion to seroreversion, while fixing standard deviation at 50 days (supplementary method). The daily seroprevalence (*P*_*t*_) was calculated by dividing *S*_*t*_ by the population (8.3 million for New York City and 3.7 million for Connecticut). We compared the estimated *P*_*t*_ with the reported seroprevalence in each round of the CDC commercial laboratory serosurvey, and calculated the log-likelihood assuming the binomial distribution, which was then used in the MCMC analysis for parameter estimation (supplementary methods).

**Figure 1.**
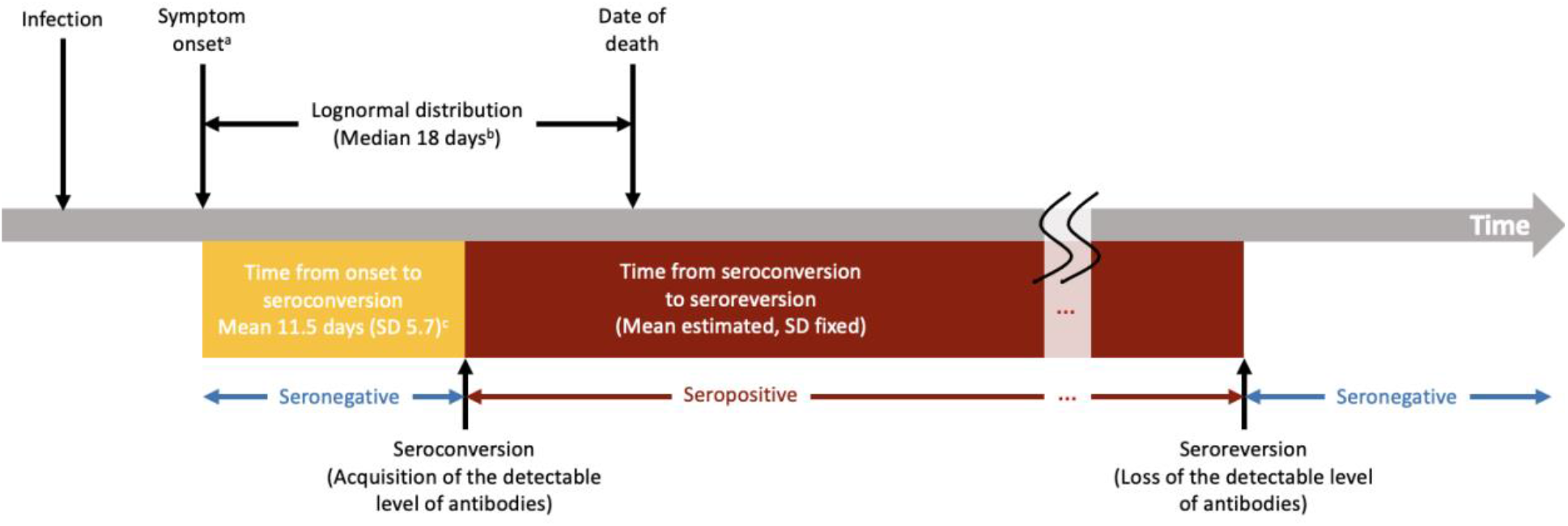
Structure of the analytic framework. ^a^Onset of infectiousness for asymptomatic cases; ^b^Data from Georgia Department of Public Health; ^c^Iyer *et al*. medRxiv 2020. Abbreviations: SD, standard deviation.

Given an estimated model from above, the number of individuals who seroconverted on day *t* (*C*_*t*_) may be calculated as follows:

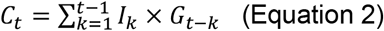

where *G* is the cumulative density function of the aforementioned Weibull distribution for time from symptom onset to seroconversion. *G*_*t*−*k*_ represents the probability that infected individuals on day *k* (*I*_*k*_) had seroconverted by day *t*. We calculated the cumulative incidence on day *t* by dividing the cumulative sum of *C*_*t*_ by the population. We also calculated a cumulative case ascertainment ratio (*A*_*t*_) as follows:

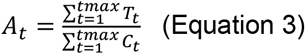

where *T*_*t*_ is the number of documented cases on day *t* (supplementary methods). The ascertainment bias was calculated as 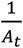.

The median of the posterior samples was reported as a point estimate, and 2.5th and 97.5th percentiles were reported as 95% credible intervals (CrIs). All analyses were performed with R (Vienna, Austria). The code can be found in the following github repository: https://github.com/lopmanlab/SARS-CoV-2_CumInc_WaningAntibodies.

### Sensitivity analysis

We performed sensitivity analyses to evaluate how sensitive our results were to different assumptions. We changed the fixed value of the standard deviation for time from seroconversion to seroreversion from 50 days to 20 and 70 days. We selected these values based on previous findings.^20^ We also relaxed the assumption of the constant IFR. It has been reported that the IFR and case fatality ratio for COVID-19 declined over time in New York City^21^ and in other countries.^20^ Therefore, we assumed that the IFR decreased by 5% per week from mid-March to the end of July 2020, reflecting findings reported by the previous studies,^21^ and the model estimated the IFR in mid-March (before the decline started). Lastly, we used data from Wuhan, China to inform delay between symptom onset and death,^22^ instead of the Georgia data to evaluate the impact of variation in this distribution on parameter inference.^23^

### Ethical considerations

Seroprevalence data, mortality data, and case data in New York City and Connecticut were publicly available, deidentified, and aggregated. The Georgia Department of Public Health Institutional Review Board (IRB) has determined that the project is exempt from the requirement for IRB review and approval.

## Results

### Estimated timeline of seroreversion, IFR, and case ascertainment ratio

The average time from seroconversion to seroreversion was estimated to be 4.0 months (95% CrI: 3.6-4.6 months) using the New York City data and 3.0 months (95% CrI: 2.3-4.1 months) using the Connecticut data with the standard deviation fixed at 50 days (Figure 2). More than 85% of the infected individuals were estimated to become seronegative due to waning antibodies within six months after seroconversion (Table 1).

**Table 1.**
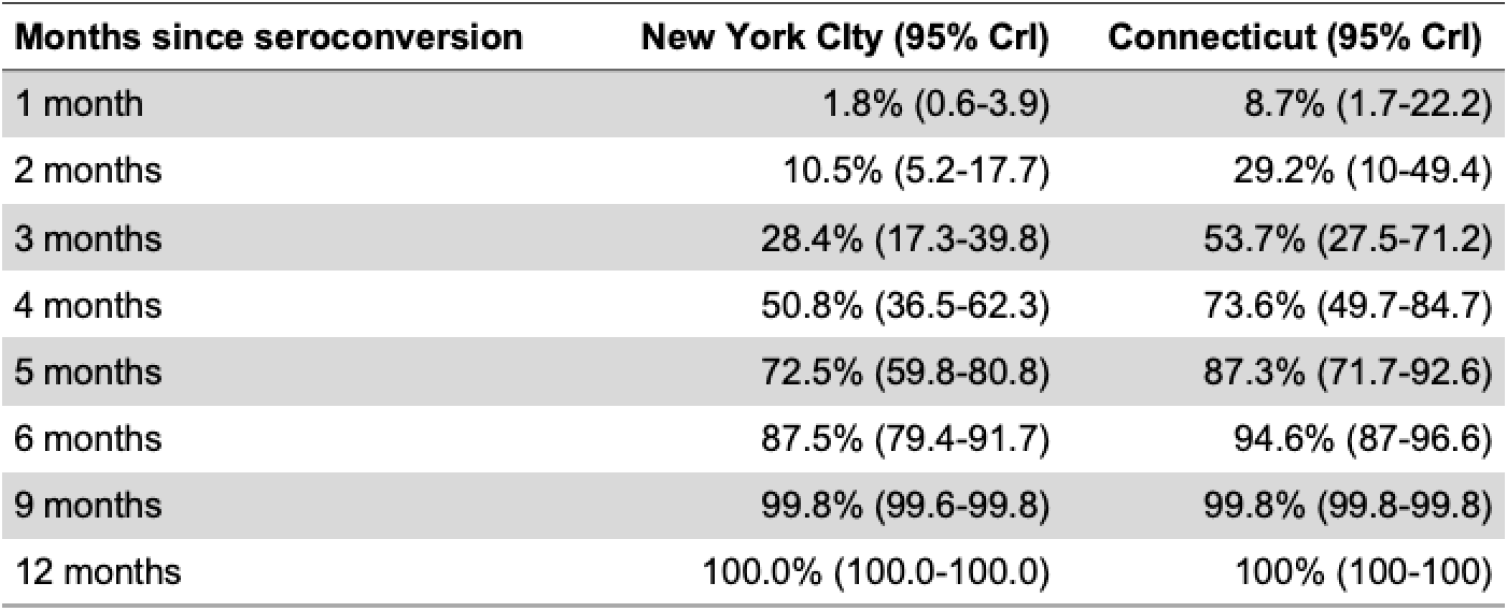
Percentage of infected individuals whose antibody level has declined and become undetectable by serological assays within *t* months since seroconversion. The mean of the Weibull distribution for time from seroconversion to seroreversion was estimated while fixing the standard deviation at 50 days. Abbreviation: CrI, credible interval.

| **Months since seroconversion** | **New York City (95% CrI)** | **Connecticut (95% CrI)** |
| --- | --- | --- |
| 1 month | 1.8% (0.6-3.9) | 8.7% (1.7-22.2) |
| 2 months | 10.5% (5.2-17.7) | 29.2% (10-49.4) |
| 3 months | 28.4% (17.3-39.8) | 53.7% (27.5-71.2) |
| 4 months | 50.8% (36.5-62.3) | 73.6% (49.7-84.7) |
| 5 months | 72.5% (59.8-80.8) | 87.3% (71.7-92.6) |
| 6 months | 87.5% (79.4-91.7) | 94.6% (87-96.6) |
| 9 months | 99.8% (99.6-99.8) | 99.8% (99.8-99.8) |
| 12 months | 100.0% (100.0-100.0) | 100% (100-100) |

**Figure 2.**
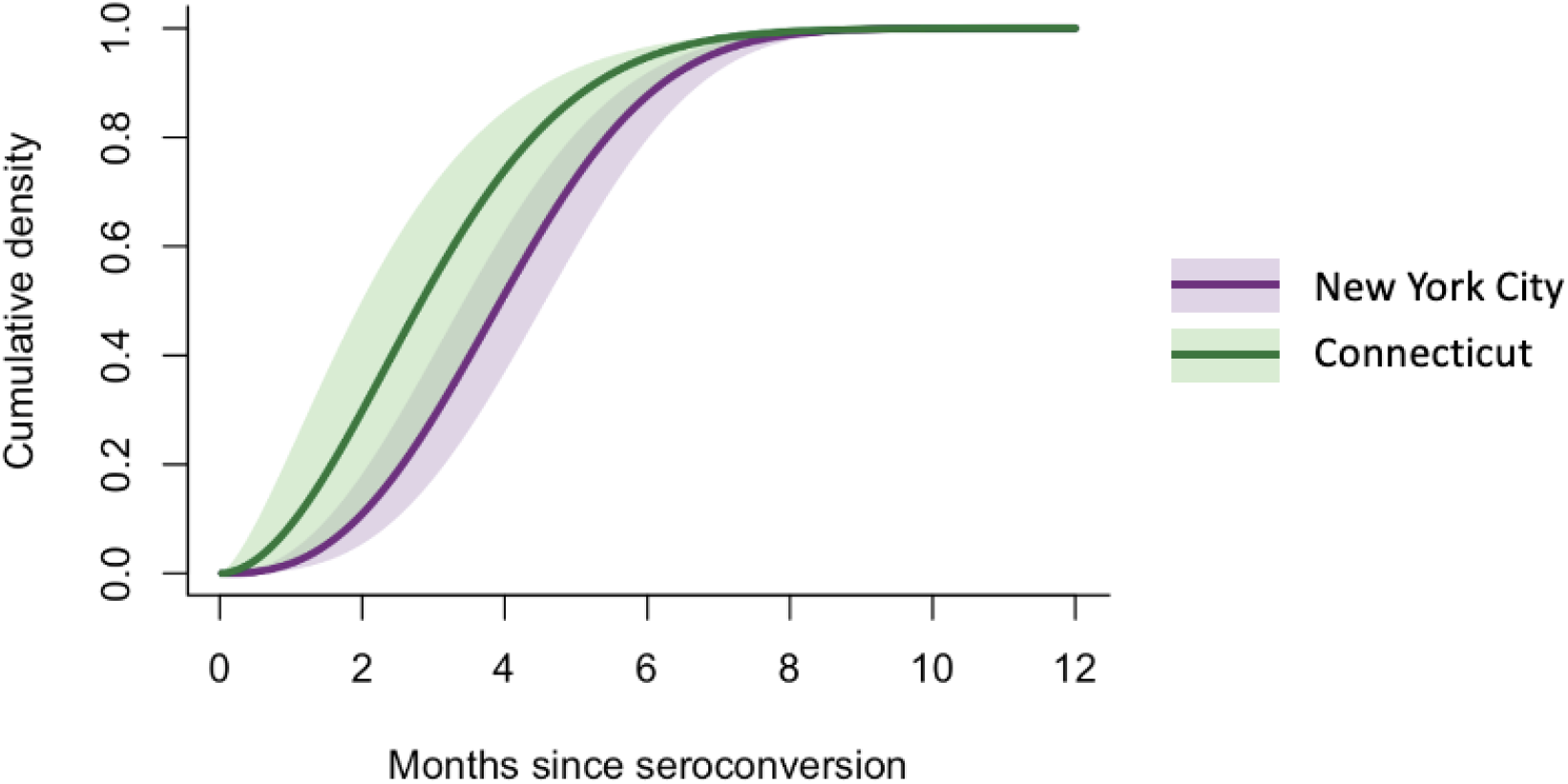
Cumulative density function for the estimated Weibull distribution for time from seroconversion to seroreversion (New York City and Connecticut). Lines and shaded areas represent the posterior median and 95% credible intervals, respectively.

The IFR was estimated to be 1.1% (95% CrI: 1.0-1.2%) for New York City and 1.4% (95% CrI: 1.1-1.7%) for Connecticut. The estimated case ascertainment ratio increased rapidly in the early phase of the pandemic and continued to gradually increase from May to October in both sites (Figure 3). The ascertainment ratio was estimated to reach 13% (95% CrI: 12-14%) in New York City and 18% (95% CrI: 14-23%) in Connecticut at the end of September 2020, suggesting that the number of estimated infections was 7.7 and 5.6 times greater than the number of documented cases in New York City and Connecticut, respectively.

**Figure 3.**
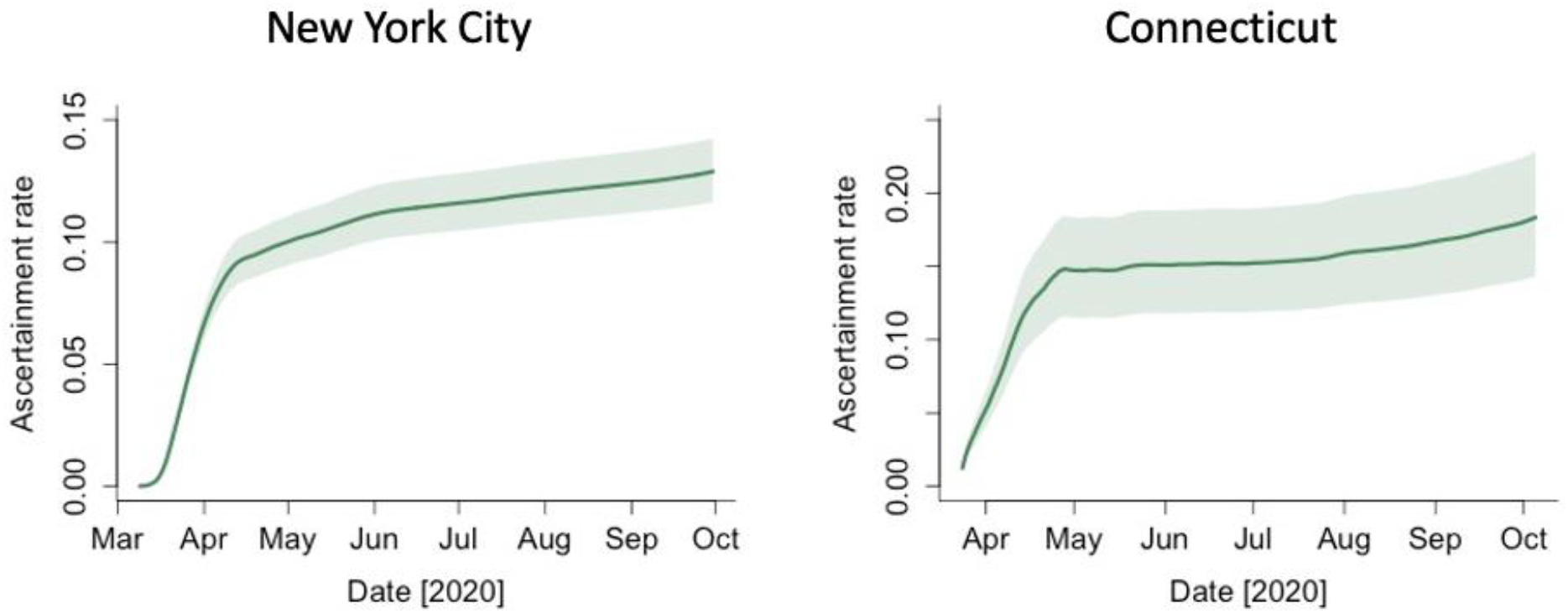
Estimated case ascertainment ratio in New York City and Connecticut in 2020. The case ascertainment ratio was calculated in Equation 3. Lines represent the 50th percentile of the posterior distributions, and shaded areas represent 95% credible intervals.

### Estimated daily seroprevalence and cumulative incidence

In both New York City and Connecticut, the estimated daily seroprevalence decreased over time after the first wave of COVID-19, diverging from the estimated cumulative incidence adjusted for waning antibodies (Figure 4). The cumulative incidence was estimated to reach 26.8% (95% CrI: 24.2-29.7%) in New York City and 8.8% (95% CrI: 7.1-11.3%) in Connecticut at the end of September 2020. In contrast, the estimated daily seroprevalence peaked at 22.1% (95% CrI: 20.5-23.6%) in mid-May and declined to 4.9% (95% CrI: 3.9-6.9%) by the end of September in New York City. In Connecticut, the estimated daily seroprevalence peaked at 6.1% (95% CrI: 5.3-7.1%) at the end of May and early June and declined to1.3% (95% CrI: 0.9-2.0%) by the end of September.

**Figure 4.**
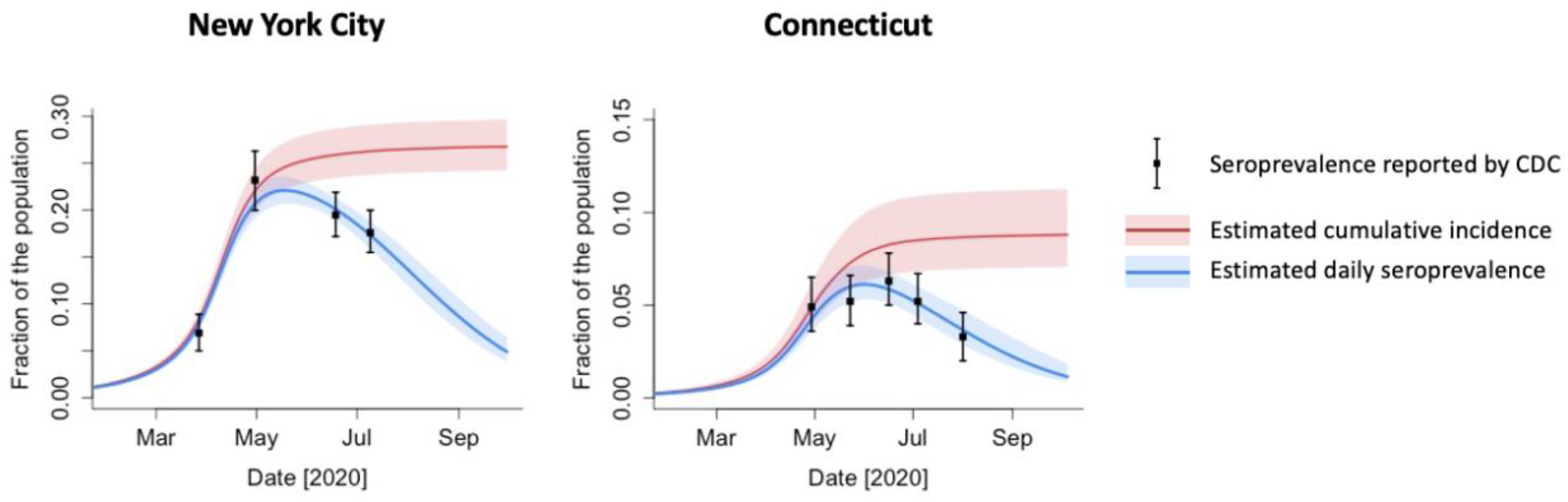
Estimated daily seroprevalence and cumulative incidence of SARS-CoV-2 infection in New York City and Connecticut in 2020. Black dots and bars represent the point estimates and 95% confidence intervals for the seroprevalence reported by CDC.^12^ Abbreviations: SARS-CoV-2, severe acute respiratory syndrome coronavirus 2; CDC, Centers for Disease Control and Prevention.

### Sensitivity analysis

We ran the analysis with different values of the standard deviation for time from seroconversion to seroreversion (20, 50, and 70 days). The estimated IFR and average time from seroconversion to seroreversion were robust to this variation for Connecticut, but the estimated mean time increased as the standard deviation increased for the New York City data (Table S2). Although changes in the standard deviation had small effects on the estimated cumulative incidence, the speed of decline in the daily seroprevalence after the peak varied by standard deviation (Figure S2). When the standard deviation was 20 days, the daily seroprevalence declined rapidly after the peak and became 1.2% (95% CrI: 0.9-2.1%) at the end of September, while it declined more slowly with the standard deviation 70 days (7.4% (95% CrI: 6.1-8.7%) at the end of September).

Under the assumption of the decreasing IFR (Figure S3), the average time from seroconversion to seroreversion was estimated to be 4.0 months (95% CrI: 3.5-4.8 months) with the standard deviation 50 days in New York City, which was consistent with the result using the constant IFR (Table S2). The estimated IFR in mid-March was 2.9% (95% CrI: 2.6-3.3%), which automatically decreased by 5% per week and reached 1.1% (95% CrI: 0.9-1.2%) at the end of July (Figure S3). The average IFR from March to September was 1.4% (95% CrI: 1.2-1.6%). The daily seroprevalence and cumulative incidence did not appreciably change between the models using the constant IFR vs. decreasing IFR (Table S2). We also used data from Wuhan, China to account for variation in the delay between symptom onset and death (Figure S4), and found that results were robust to this change (Table S2).

## Discussion

A reliable estimate of the cumulative number of people who have been infected with SARS-CoV-2 is critical for understanding and, ultimately, controlling the COVID-19 pandemic. Measuring severity (in the form of the IFR), monitoring progress towards a herd immunity threshold, and predicting the impact of vaccination all depend on a robust estimate of cumulative incidence of infection. Given evidence that anti-SARS-CoV-2 antibodies wane below the detection limit among a substantial portion of the population, we developed an analytical framework to estimate cumulative incidence of infection from cross-sectional serological surveys. First, our model was able to capture the observed declines in seroprevalence after the first wave of COVID-19 in the spring of 2020 in New York City and Connecticut. Second, we were able to estimate that the cumulative incidence was 26.8% (95% CrI: 24.2-29.7%) in New York City and 8.8% (95% CrI: 7.1-11.3%) in Connecticut by the end of September 2020, which was greater than the peak daily seroprevalence of 22.1% (95% CrI: 20.5-23.6%) in mid-May in New York City and 6.1% (95% CrI: 5.3-7.1%) at the end of May in Connecticut. Cumulative incidence could be underestimated by cross-sectional serosurveys only a few months after the first cases of SARS-CoV-2 in a population, because individuals no longer have a detectable level of antibodies 3-4 months after seroconversion on average. Taken together, our findings suggest that cumulative incidence of SARS-CoV-2 should not be estimated directly from cross-sectional serology data, especially at later stages of an outbreak. Rather, cumulative incidence should be adjusted given impacts of waning of detectable antibodies in serological assays.

The timeline of seroconversion and seroreversion should ideally be determined by individual-level longitudinal studies that follow up each infected individual multiple times, as frequently as possible, for a long enough period to observe seroreversion. The timeline for seroreversion after SARS-CoV-2 infection reported by the longitudinal studies published to date varies. Some studies have reported rapid waning of IgG, with substantial attrition of the seropositive population in as little as 60 days,^7,24^ while others reported that antibodies remained above the detectable threshold for at least 82 days after symptom onset^25^ or 120 days after qPCR diagnosis of SARS-CoV-2.^26^ These differences are likely due to limitations or heterogeneity of the studies reported to date, including short follow-up times, small sample sizes, and different serology testing methods and analytic sensitivities.^27^ Most of the longitudinal studies published to date have followed participants for 14-150 days after symptom onset or baseline visits, which is not long enough to understand the complete timeline of seroreversion for all participants, particularly for IgG.^7,8,19,20,24,25^ The variation in clinical and demographic characteristics and severity of infection of participants in each study has also likely influenced these different findings.^16,17,20,24,28^ Infected individuals with mild or no symptoms who may exhibit lower titer and shorter time to seroreversion are often not included in these longitudinal studies. Therefore, assessing the timeline using population-level data, such as cross-sectional serology data and mortality data, is a viable alternative approach. Using our framework, infected individuals were estimated to remain seropositive for about 3-4 months on average. The average duration estimated by the New York City data and Connecticut data were mostly in agreement, with overlapping credible intervals. Differences in these average durations could be attributable to differences in demographic and clinical characteristics of infected individuals and differences in the testing and reporting practices. It is also important to note that the timeline of seroconversion and seroreversion is dependent on serological assays and targeted immunoglobulins that may have different thresholds to define seropositivity.

We estimated that the IFR was 1.1% (95% CrI: 1.0-1.2%) in New York City, which was consistent with estimates from the routine care group at Mount Sinai Hospital in New York City.^29^ Yang *et al*. estimated the time-varying IFR for New York City (1.39%; 95% CrI: 1.04-1.77),^21^ matching with our estimate under the assumption of decreasing IFR (1.4% (95% CI: 1.2-1.6%) on average between March and September 2020). Although the sensitivity analyses did not change our central conclusions, we note that different values of the standard deviation for time from seroconversion to seroreversion changed the estimated seroprevalence in the later phase of the study period (August and September 2020; Figure 4 and Figure S2). We ran the model with standard deviation 20, 50, and 70 days, because the estimated standard deviation for time from seroconversion to seroreversion for IgA and IgM was approximately 50 and 25 days, respectively.^20^ Our standard deviation could be larger than those estimated for small study populations, as we used population-level data that likely have greater variation in demographic and clinical characteristics. Analyzing seroprevalence data in August and after could provide critical information on how population-level seroprevalence declines over time and would enable better estimates of the timeline of antibody waning.

The data used in this study have certain limitations. The samples collected for the CDC seroprevalence data may not be representative of the general population, as Havers *et al*. discussed.^6^ The data on the date of symptom onset among cases who died in Georgia may be incorrectly recorded and subject to recall bias as they are largely self-reported. Changes in testing and reporting practice over time likely affected the number of reported deaths, although mortality data are less sensitive to these changes compared to case data. We conducted the sensitivity analysis with a different source of data on time from onset to death and considered time-varying IFR instead of constant IFR, and found that results were robust to these changes (Table S2).

Detection of antibodies by serological assays may not correlate to protection from reinfection or disease, and thus, time from seroconversion to seroreversion estimated in our study is not necessarily equivalent to the duration of protective immunity. Several studies have noted strong correlation between results from assays based on antibody binding (such as ELISA) and neutralization testing,^16,20,25^ which may allow protective immunity to be inferred from simpler serologic tests once more comprehensive longitudinal data sets are available. Moreover, T cell immunity may make an important contribution to protective immunity but is not assessed by serologic methods.^30^ Therefore, additional studies are needed to define immunologic determinants of SARS-CoV-2 protection against reinfection and severe disease.

Our findings suggest that the cumulative incidence estimated from serology data needs to be adjusted for seroreversion. We intend to apply this framework to other seroprevalence studies^31^ and we suggest others conducting serological surveys consider doing so as well. Our framework could readily be applied to other data to estimate the duration of seroreversion, IFR, ascertainment ratios, daily seroprevalence, and cumulative incidence.

## Data Availability

We used publicly available, deidentified, aggregate data downloaded from the government websites cited in the paper.

## Acknowledgements

The authors thank Dr. Laura Edison from the Georgia Department of Public Health for sharing the data. We also thank Dr. Manish Patel from the Centers for Disease Control and Prevention, Carly Adams, Avnika B Amin, and Dr. Julia Baker from Emory University for providing feedback on this study.

## Source of funding

This study was supported by the US National Science Foundation [grant 2032082 to JSW and grant 2032084 to BAL] and the US National Institute of Allergy and Infectious Diseases [3R01AI143875-02S1 to PSS and AJS].

## Conflicts of interest

BAL reports personal fees from Takeda Pharmaceutical, personal fees from CDC Foundation, and personal fees from Hall Booth Smith, P.C., outside the submitted work. Other authors do not have conflicts of interest.

## Supplementary materials

### Supplementary methods

#### More details on the population-level cross-sectional seroprevalence data

For New York City, the catchment area covered Manhattan, Bronx, Queens, Kings, and Nassau Counties in the first round (catchment population: 9.26 million), with Suffolk, Westchester, and Richmond Counties added in the second, third, and fourth round (total catchment population: 12.2 million). Because the county-specific serology data were not available, we assumed that the seroprevalence was similar across the different catchment areas in the New York City metro area. For Connecticut, the catchment area included all counties in the state.

#### More details on the mortality data and case data

The mortality data for New York City covered Bronx, Kings, Manhattan. Queens, Richmond Counties. The mortality data covered all counties in the state. For Connecticut, we calculated a 7-day rolling average, because there were no data (i.e., zero deaths reported) on weekends. For New York City, we used the reported daily counts.

For the number of documented cases (*T*_*t*_ in Equation 3), we used the number of individuals with positive PCR and/or antigen tests in New York City. In Connecticut, *T*_*t*_ denotes the number of total cases (sum of confirmed and probable cases), because confirmed cases and probable cases were not reported separately before May 31, 2020.

#### Total number of SARS-CoV-2 infections

To estimate the total number of SARS-CoV-2 infections (*I*_*t*_) and the IFR, we first estimated the date of symptom onset for fatal cases, by fitting gamma, Weibull, and lognormal distributions to the observed data on time from symptom onset to death for 6,999 COVID-19 associated deaths in Georgia. The best fitted distribution was selected based on goodness-of-fit tests (Cramer-von Mises, Kolmogorov-Smirnov and Anderson-Darling statistics), using the fitdistrplus R package.^32^ Among those who died on day *t* (*d*_*t*_), we calculated the number of individuals who developed symptoms on day *t-k* (*o*_*t*−*k*_) as follows:

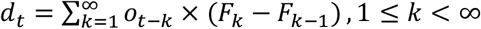

where *F* is the cumulative density function of the best fitted distribution for time from onset to death, conditional upon fatality. We then calculated the total number of infections on day *t* (*I*_*t*_) as follows:

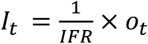

This framework assumes that the duration of incubation period is approximately equal to the duration of latent period, and therefore, day *t* represents the onset of infectiousness for asymptomatic individuals.

#### Markov chain Monte Carlo (MCMC) analysis

In the MCMC analysis, we obtained at least 50,000 posterior samples after a burn-in period of 20,000 iterations. Model convergence was assessed based on trace plots of posterior samples and the Geweke diagnostic test. We fixed the standard deviation for the Weibull distribution for time from seroconversion to seroreversion in the analysis, as parameters were not identifiable otherwise.

## Supplementary tables

**Supplementary Table S1.** Date of the CDC longitudinal commercial laboratory seroprevalence surveys in New York City metro area and Connecticut in 2020.

| Rounds of the survey | Sampling period (number of samples collected) |  |
| --- | --- | --- |
|  | New York City metro area | Connecticut |
| Round 1 | March 23 - April 1 (n=2482) | April 26 - May 3 (n=1431) |
| Round 2 | April 25 - May 6 (n=1116) | May 21- May 26 (n=1800) |
| Round 3 | June 15 - June 21 (n=1581) | June 15 - June 17 (n=1798) |
| Round 4 | July 7 - July 11 (n=1602) | July 3 - July 6 (n=1802) |
| Round 5 | NA* | July 30 - August 3 (n=992) |

\*From August, the CDC commercial laboratory survey was expanded to cover other parts of New York State. Results were reported at state level, and data specific for the New York City metro area was not available. We therefore did not include the Round 5 data in this study. Abbreviation: CDC, Centers for Disease Control and Prevention.

**Supplementary Table S2.**
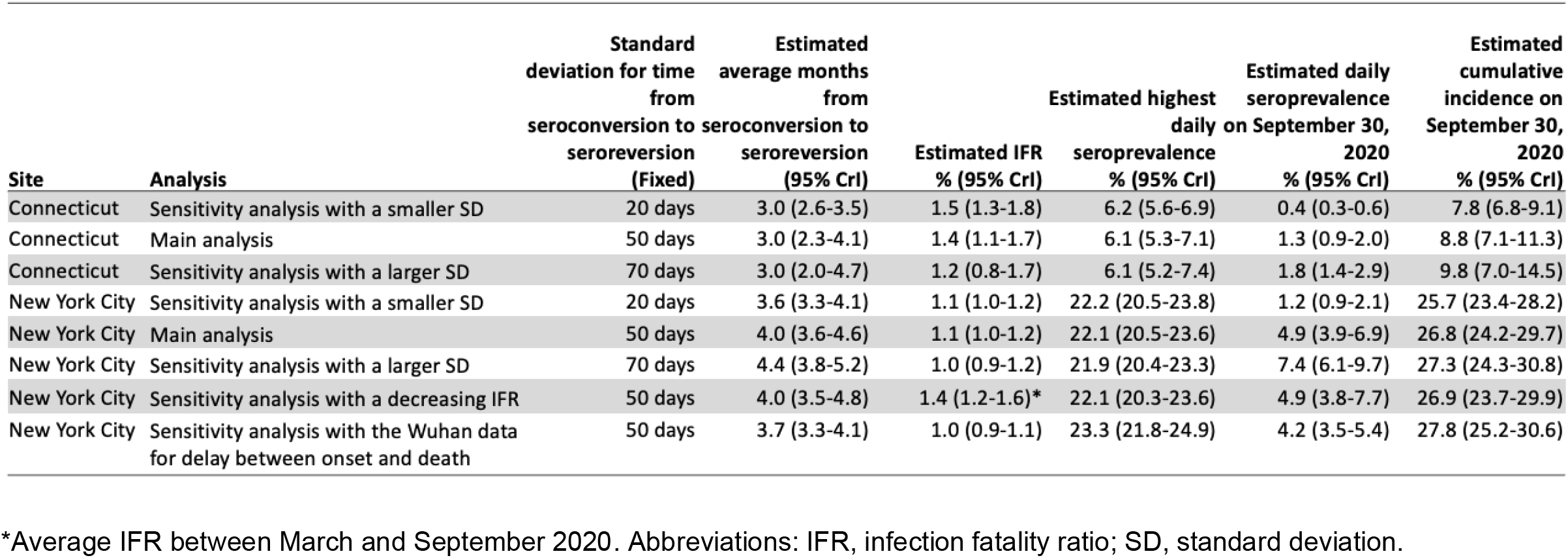
Results of the main analysis and sensitivity analysis.

## Supplementary figures

**Figure S1.**
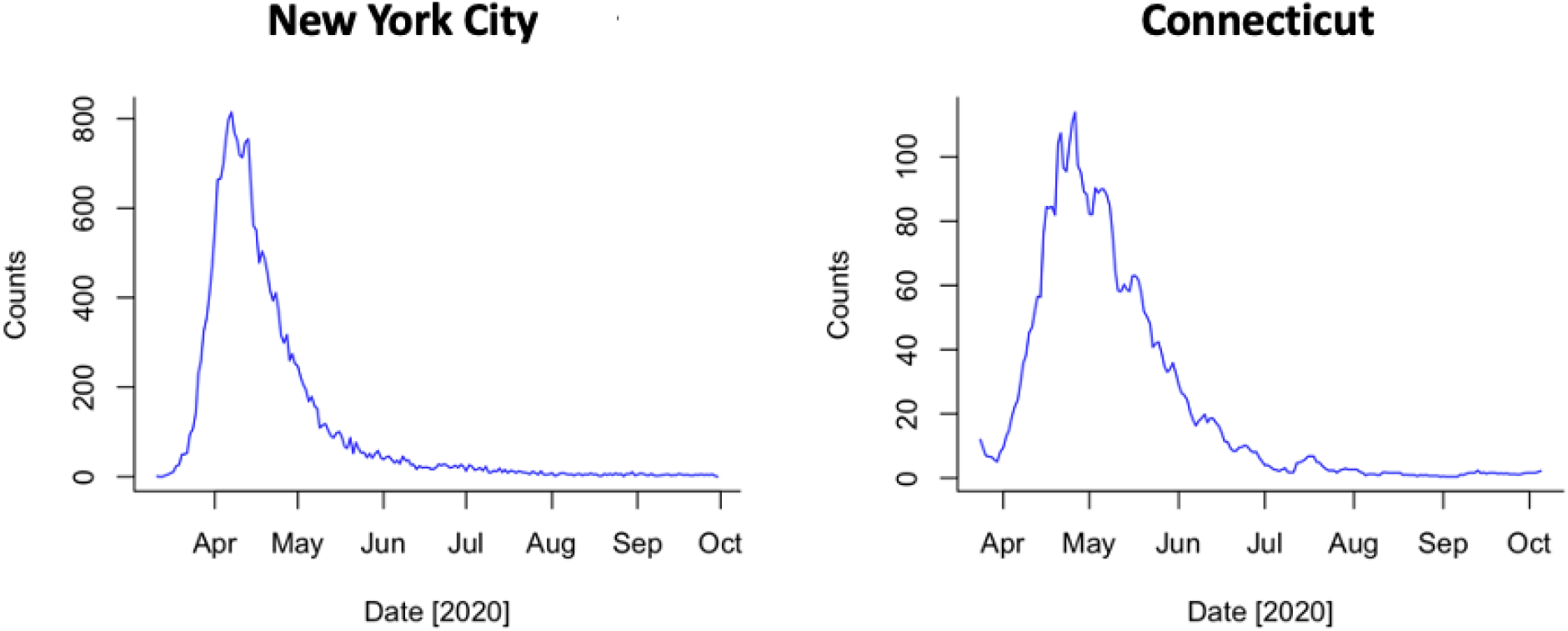
The number of reported deaths in New York City and Connecticut in 2020. A seven-day rolling average is shown for Connecticut.

**Figure S2.**
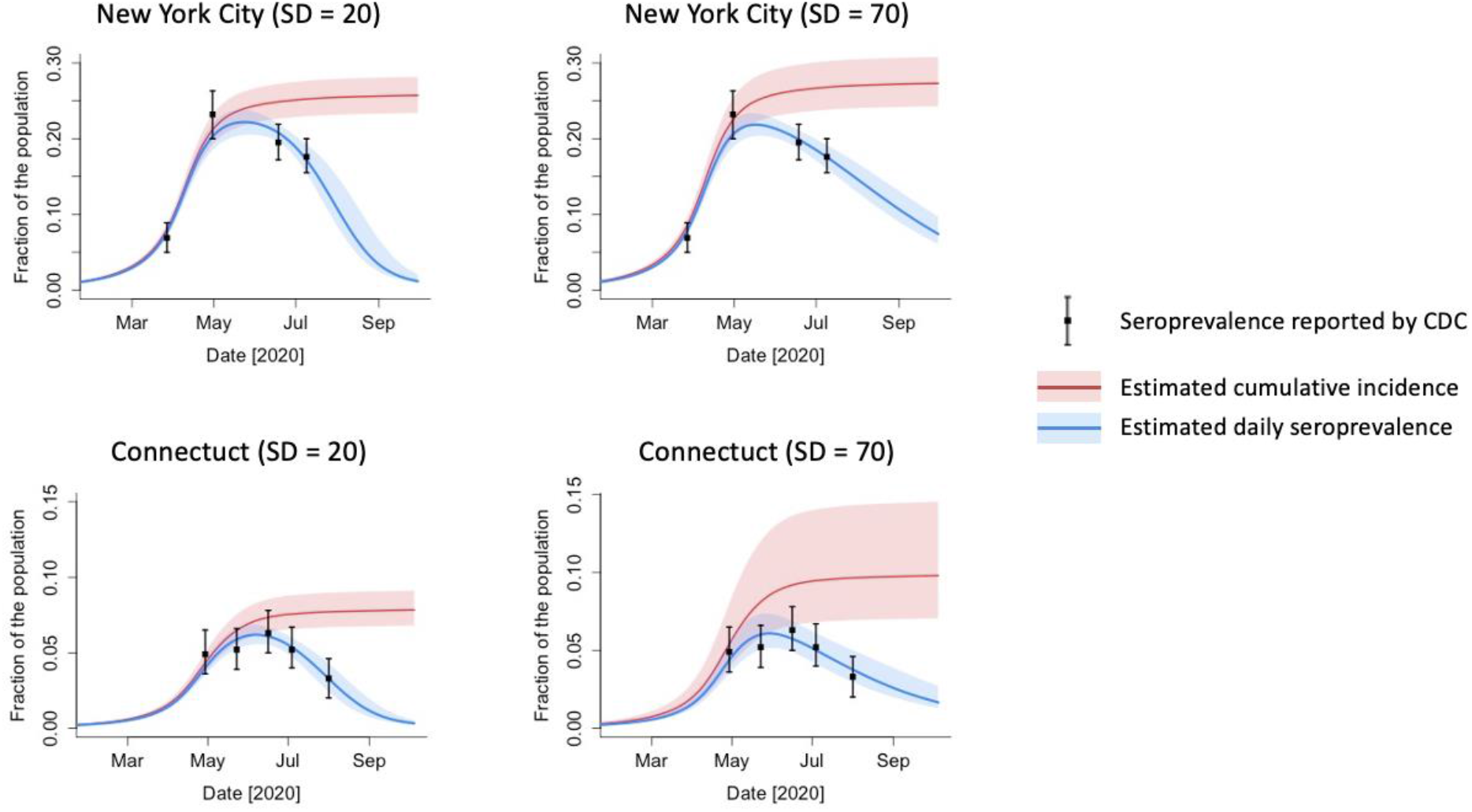
Estimated daily seroprevalence and cumulative incidence of SARS-CoV-2 infection in New York City and Connecticut in 2020 with different values of the standard deviation for time from seroconversion to seroreversion. Black dots and bars represent are the point estimates and 95% confidence intervals for the seroprevalence reported by CDC.^12^ Abbreviations: SARS-CoV-2, severe acute respiratory syndrome coronavirus 2; SD, standard deviation; CDC, Centers for Disease Control and Prevention.

**Figure S3.**
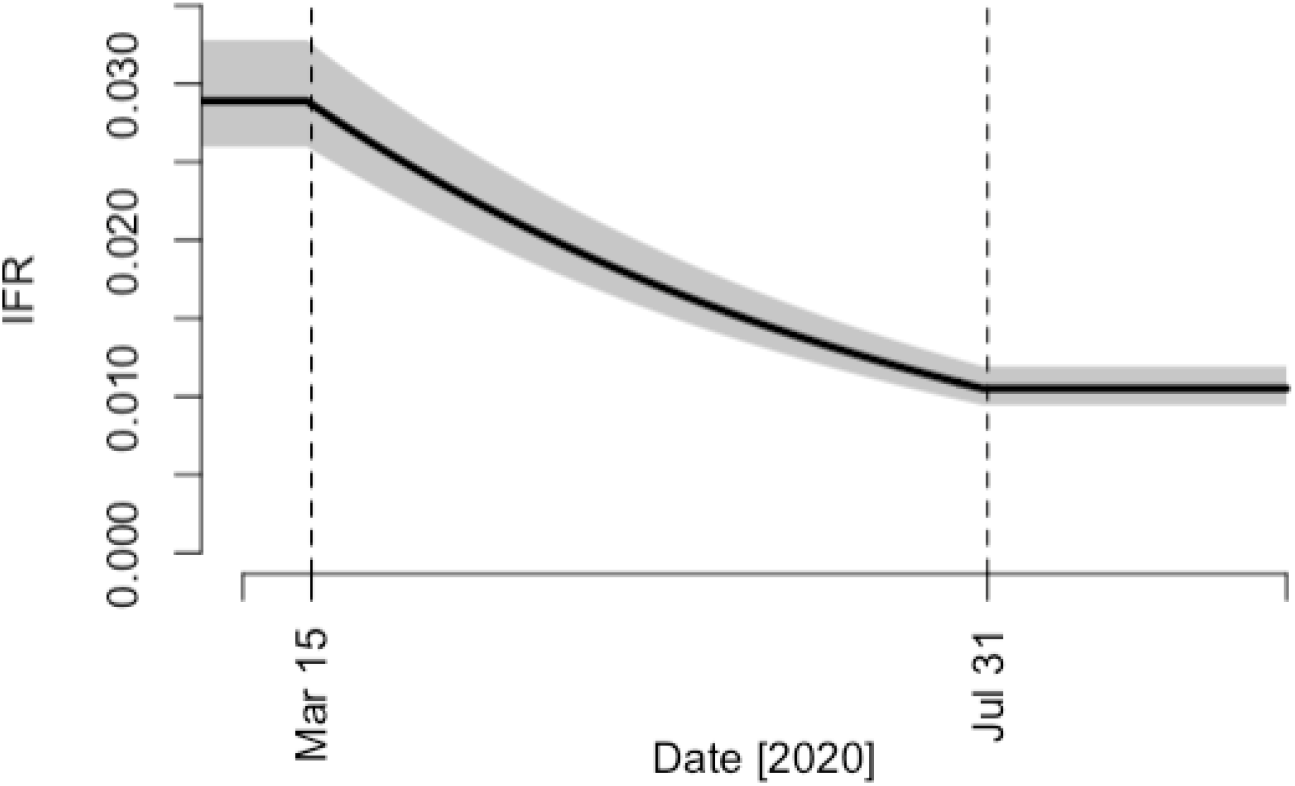
Estimated infection fatality ratio in New York City under the assumption that it decreased by 5% per week from March 15 to July 31 in 2020. A line represents the median of posterior samples and a shaded area represents the 95% credible interval. Abbreviation: IFR, infection fatality ratio.

**Figure S4.**
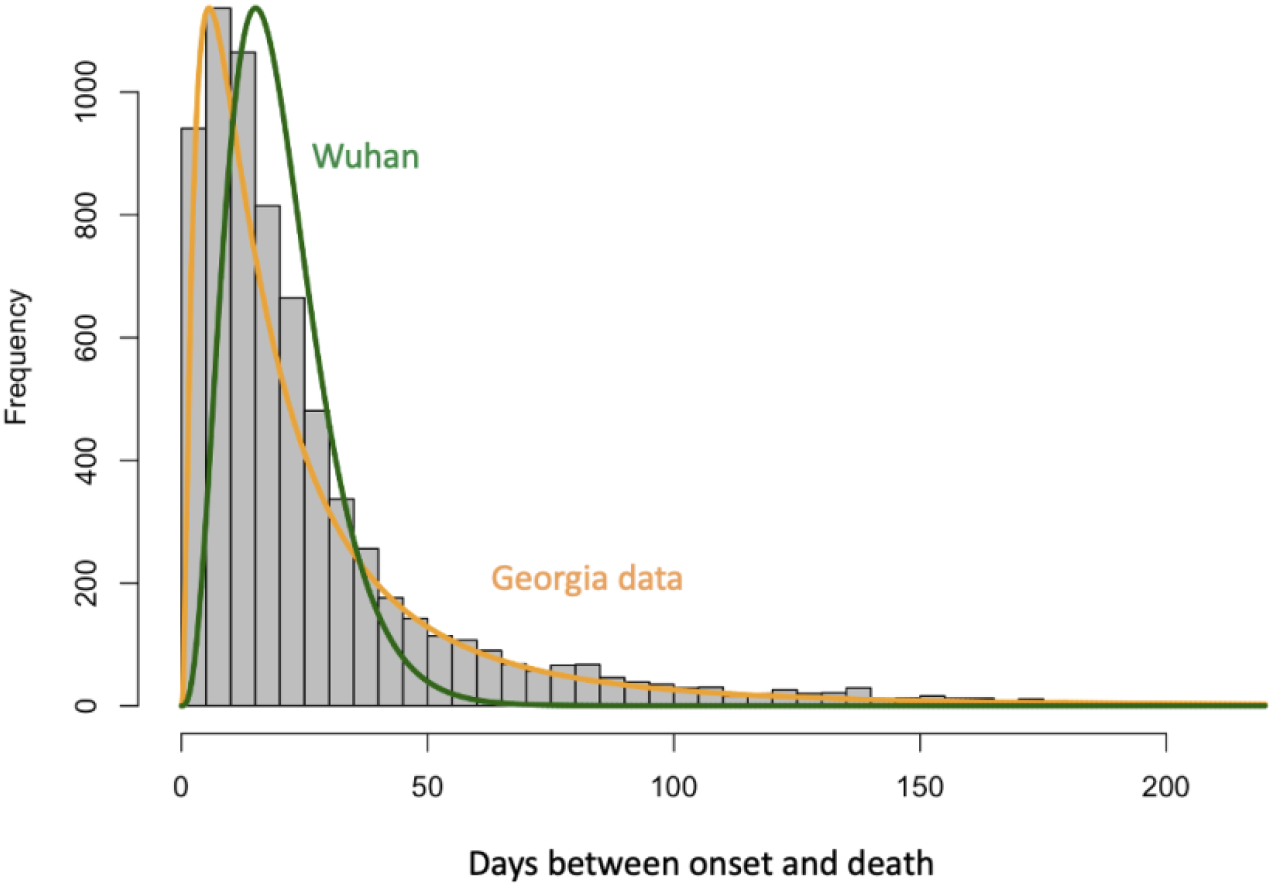
Time from symptom onset to death for COVID-19 fatal cases in Georgia, USA and Wuhan, China. The histogram represents the raw data on time from onset to death in Georgia, and an orange line is its best fitted distribution. A green line represents the best fitted distribution to the data from Wuhan, China.^22^ The original data from Wuhan, China can be found in the previous study and is not reproduced here.^22^ Abbreviations: COVID-19: coronavirus disease 2019.

## Notes

### Author Declarations

Georgia Department of Public Health Institutional Review Board

